# Enhancing epidemiological investigation of nosocomial SARS-CoV-2 infection with whole genome sequencing: A retrospective cohort study across four hospitals in the UK

**DOI:** 10.1101/2021.06.28.21259028

**Authors:** Sheila F Lumley, Bede Constantinides, Nicholas Sanderson, Gillian Rodger, Teresa L Street, Jeremy Swann, Kevin K Chau, Denise O’Donnell, Fiona Warren, Sarah Hoosdally, OUH Microbiology laboratory, OUH Infection Prevention and Control team, Anne-Marie O’Donnell, Timothy M Walker, Nicole E Stoesser, Lisa Butcher, Tim EA Peto, Derrick W Crook, Katie Jeffery, Philippa C Matthews, David W Eyre

## Abstract

**Background:** Despite robust efforts, patients and staff acquire SARS-CoV-2 infection in hospitals. In this retrospective cohort study, we investigated whether whole-genome sequencing (WGS) could enhance the epidemiological investigation of healthcare-associated SARS-CoV-2 acquisition.

**Methods and findings:** From 17-November-2020 to 5-January-2021, 803 inpatients and 329 staff were diagnosed with SARS-CoV-2 infection across four teaching hospitals in Oxfordshire, UK. We classified cases according to epidemiological definitions, sought epidemiological evidence of a potential source for each nosocomial infection, and evaluated if epidemiologically-linked cases had genomic evidence supporting transmission. We compared epidemiological and genomic outbreak identification.

Using national epidemiological definitions, 109/803 (14%) inpatient infections were classified as definite/probable nosocomial, 615 (77%) as community-acquired and 79 (10%) as indeterminate. There was strong epidemiological evidence to support definite/probable cases as nosocomial: 107/109 (98%) had a prior-negative PCR in the same hospital stay before testing positive, and 101(93%) shared time and space with known infected patients/staff. Many indeterminate cases were likely infected in hospital: 53/79 (67%) had a prior-negative PCR and 75 (95%) contact with a potential source. 89/615 (11% of all 803 patients) with apparent community-onset had a recent hospital exposure.

WGS highlighted SARS-CoV-2 is mainly imported into hospitals: within 764 samples sequenced 607 genomic clusters were identified (>1 SNP distinct). Only 43/607 (7%) clusters contained evidence of onward transmission (subsequent cases within ≤1 SNP). 20/21 epidemiologically-identified outbreaks contained multiple genomic introductions. Most (80%) nosocomial acquisition occurred in rapid super-spreading events in settings with a mix of COVID-19 and non-COVID-19 patients. Hospitals not routinely admitting COVID-19 patients had low rates of transmission. Undiagnosed/unsequenced individuals prevent genomic data from excluding nosocomial acquisition.

**Conclusions:** Our findings suggest current surveillance definitions underestimate nosocomial acquisition and reveal most nosocomial transmission occurs from a relatively limited number of highly infectious individuals.

## Introduction

Limiting acquisition of SARS-CoV-2 by patients and staff in hospitals is an infection prevention and control (IPC) priority. Despite robust efforts, both patients and staff are infected in hospitals; 10-40% of hospital-diagnosed COVID-19 cases are thought to have been acquired in hospital, with 8700 deaths following nosocomial infection reported in the UK,[1–4] and higher rates of seroconversion are reported in healthcare workers compared to the general population.[5,6]

Distinguishing which patients have acquired infection in hospital allows potential transmission events to be investigated. Epidemiological rules are frequently used for nosocomial classification and outbreak investigation, using spatial and temporal patient data to make assumptions about acquisition and transmission. However, such rules may exclude plausible transmission and leave uncertainty around the source of individual infections. SARS-CoV-2 whole-genome sequencing (WGS) has been proposed as an adjunct to assist hospital outbreak investigation. Individuals infected with identical or near-identical (≤1 SNP) viruses, are more likely to be linked in a transmission chain than those with more distantly related viruses, as demonstrated by previous retrospective studies that have utilised WGS to identify nosocomial infections and outbreaks.[7–12]

We investigated whether sequencing could enhance epidemiological investigation of healthcare-associated SARS-CoV-2 acquisition in two areas: i) confirming/excluding nosocomial acquisition and ii) understanding the role of outbreaks in nosocomial acquisition. We highlight the benefits and pitfalls of this approach, to help guide local practice in individual centres.

## Methods

### Study design, setting and participants

The Oxford University Hospitals NHS Foundation Trust comprises four hospitals with ∼1100 beds (mostly in 4-bed bays within wards of 20-30 beds) and ∼13,500 staff. The four hospitals are presented as “A”, a large acute hospital admitting both COVID-19 and non-COVID-19 patients; “B”, a smaller general hospital admitting both COVID-19 and non-COVID-19 patients; “C”, a hospital focused predominantly on cancer care; and “D”, a largely elective orthopedic hospital, with C and D not routinely admitting COVID-19 patients. Ward admission and discharge dates were available for all patients from 14 days before the first positive PCR, and the work location for those staff working exclusively or predominantly on a single ward. Public Health England (PHE) guidance for COVID-19 IPC was followed throughout the study, including the use of patient pathways, personal protective equipment (PPE), symptomatic and asymptomatic staff and patient testing (summarised in Supplement).[13]

Infections in patients and hospital staff were detected by symptomatic and asymptomatic SARS-CoV-2 PCR testing of combined nasal and oropharyngeal swabs by Thermo Fisher TaqPath assay (2553/2773, 92% samples) and other platforms (details in Supplement). PCR-positive samples were stored at -80℃ for WGS. Sequencing was attempted on all stored samples, regardless of cycle threshold (Ct) value, using the ARTIC LoCost protocol[14] (details in Supplement).

### Definitions

Nosocomial SARS-CoV-2 infection was defined following NHS England and NHS Improvement definitions:[15]

– Community-Onset, PCR-positive ≤2 days after hospital admission/attendance
– Hospital-Onset Indeterminate Healthcare-Associated, PCR-positive 3-7 days after hospital admission (hereafter referred to as “Indeterminate nosocomial”)
– Hospital-Onset Probable Healthcare-Associated, PCR-positive 8-14 days after admission (“Probable nosocomial”)
– Hospital-Onset Definite Healthcare-Associated, PCR-positive ≥15 days after admission (“Definite nosocomial”)

Enhanced nosocomial classification - prior negative PCR results (available as a result of admission screening, weekly ward screening and symptomatic testing) and admissions in the 14 days prior to diagnosis were used to determine whether additional support existed for nosocomial acquisition.

For the purpose of identifying plausible transmission events, indicative incubation periods were defined as 1-14 days prior to a positive PCR test.[16] Infectious periods were defined from 4 days before to 7 days after a positive PCR for patients, and 4 days before to the day of the positive PCR test for staff (reflecting that staff isolated at home for 10 days following a positive test).[17] Mean serial intervals, i.e. the duration between the symptom-onset time of in a transmission donor and recipient, have been estimated at 4-7 days, here 5 days is used.[18,19]

Individuals acquiring SARS-CoV-2 are denoted “recipients”, and those transmitting infection as “donors”. A “plausible donor” for a recipient, is identified by the donor and recipient being present on the same ward (“ward contact”), during the donor’s infectious period and the recipient’s incubation period. “Hospital contact” was defined as presence in the same hospital on the same calendar day, during the donor’s infectious period and the recipient’s incubation period.

Epidemiological outbreaks were defined following PHE guidance:[20] ≥2 cases of COVID-19 in patients or hospital staff ‘associated with the same setting’, with ≥1 case (if a patient) meeting the definition of probable/definite nosocomial infection, ending when no cases were diagnosed for 28 days. Here ‘associated with the same setting’ was defined as a ward contact.

Genomic outbreaks were defined as for epidemiological outbreaks with the additional requirement for individual viral sequences to be genomically linked. Genomically linked sequences were defined as those sharing ≤1 SNPs, an association close enough to support transmission whilst minimising over-calling of linkage in non-nosocomial cases (see Supplement). Genomic clusters were defined as for genomic outbreaks, but without the requirement for ≥1 definite/probable nosocomial case.

### Epidemiologic and genetic analysis

We initially classified all cases according to epidemiological definitions above, and then tested if there was epidemiological evidence of a potential source case for each new definite/probable/indeterminate patient and staff infection. We then evaluated how many of these epidemiologically linked cases were within ≤1 SNPs of each other, i.e. had genomic evidence to support transmission. Following this, we searched for epidemiologically defined outbreaks involving infected patients and staff. Community-onset cases were only included as part of an outbreak if they could have plausibly seeded the outbreak (i.e. their diagnosis preceded the first staff or nosocomial patient case on that ward), and not if admitted during an ongoing outbreak.

Combined epidemiological and genomic analysis was performed using R version 4.0.2 [21], with visualisation using ggplot2[22] and igraph[23] packages. Multiple sequence alignment and phylogenetic analysis were performed with MAFFT)[24]and IQTree[25] respectively; phylogenies were prepared and visualised using Treeswift[26] and Toytree[27].

### Ethics statement

Deidentified data were obtained from the Infections in Oxfordshire Research Database which has generic Research Ethics Committee, Health Research Authority and Confidentiality Advisory Group approvals (19/SC/0403, 19/CAG/0144).

### Data sharing

Study sequence data have been deposited in European Nucleotide Archive under study accession number PRJEB43319. The epidemiological datasets analysed are not publicly available as they contain personal data but are available from the Infections in Oxfordshire Research Database (https://oxfordbrc.nihr.ac.uk/research-themes-overview/antimicrobial-resistance-and-modernising-microbiology/infections-in-oxfordshire-research-database-iord/), subject to an application and research proposal meeting the ethical and governance requirements of the Database.

## Results

### 1) Overview of cohort

From 17-November-2020 to 5-January-2021, 1132 individuals (803 inpatients, with 1104 admissions, and 329 staff) were newly diagnosed with PCR-confirmed SARS-CoV-2 infection (Figure 1). The median inpatient age at diagnosis was 67 years (IQR 49-81, range 0-102), 43% were female. 188/803 (23%) inpatient infections were classified as nosocomial (definite, probable or indeterminate). Length of stay after the first positive PCR was a median of 6 days in community-onset cases vs. 8 days in nosocomial cases (Kruskal-Wallis p<0.001). All-cause mortality within 28 days of a first positive SARS-CoV-2 PCR was 14% after community-onset and 23% after nosocomial infection (p<0.001) (Table 1).

**Table 1:** Characteristics of 803 inpatients testing SARS-CoV-2 PCR positive between 17 November 2020 and 5 January 2021 at Oxford University Hospitals.

|  | All patients | Nosocomial | Community-onset | Kruskal Wallis p-value |
| --- | --- | --- | --- | --- |
| **Total number** | 803 | 188 (23%) | 615 (77%) | NA |
| **Sex**<br>Male (n; %)<br>Female (n; %) | 454 (57%)<br>349 (43%) | 109 (58%)<br>79 (42%) | 345 (56%)<br>270 (44%) | 0.65 |
| **Age (median; IQR)** | 67 (49-81) | 76 (62-86) | 63 (46-80) | <0.001 |
| **Lowest Ct (median; IQR)** | 20 (14-27) | 19 (14-28) | 20 (15-27) | 0.15 |
| **Length of stay before first SARS-CoV-2 positive PCR (median; IQR)** | 1 (1-3) | 9 (5-15) | 1 (1-1) | <0.001 |
| **Length of stay after first SARS-CoV-2 positive PCR (median; IQR)** | 6 (2-13) | 8 (4-16) | 6 (2-11) | <0.001 |
| **28 day mortality (n; %)** | 129 (16%) | 43 (23%) | 86 (14%) | <0.001 |
| **Successfully sequenced (n; %)** | 503/803 (63%) | 116/188 (62%) | 387/615 (63%) | 0.44 |

**Figure 1:**
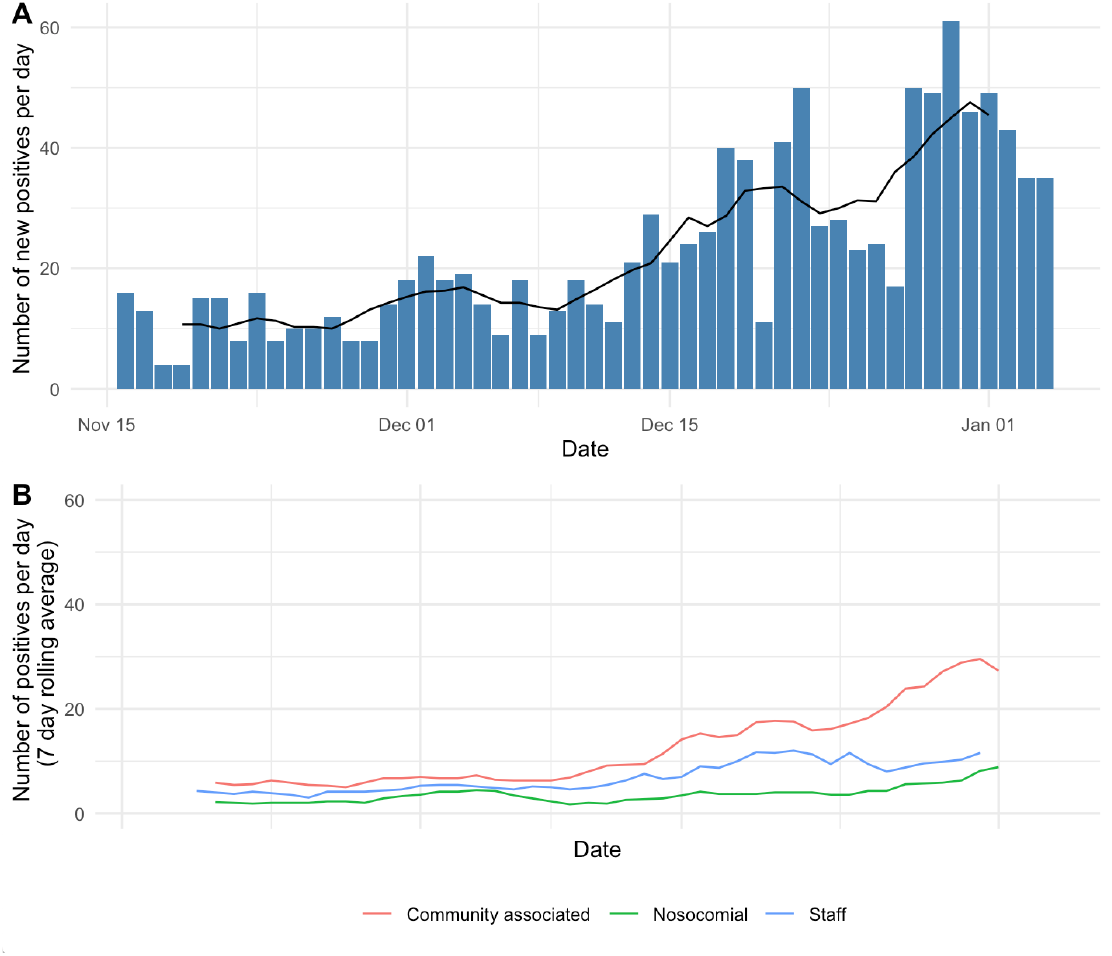
Epidemic curves showing number of new SARS-CoV-2 PCR staff and inpatient positives at Oxford University Hospitals from 17 November 2020 - 5 January 2021 based on clinical microbiology laboratory diagnosis by PCR. A) total number of staff and inpatient positives (blue bars showing new positives per day, black line shows 7 day rolling average) B) SARS-CoV-2 positives split by staff and patients (nosocomial (definite, probable and indeterminate) and community-onset according to national surveillance definitions), lines show 7 day rolling average.

Swabs from 764/1132 (67%) PCR-positive individuals were successfully sequenced, including 116/188 (62%) nosocomial cases and 261/329 (79%) staff cases (Tables S1, S2, Figure S1).

### 2) Standard epidemiological classification

#### a) Nosocomial classification

Based on standard national definitions, 188/803 (23%) inpatient infections were classified as nosocomial, subgrouped as definite (n=51), probable (n=58) or indeterminate (n=79). In the UK, patients who acquired SARS-CoV-2 infection in hospital but were discharged before testing positive are not reported as nosocomial and so are not accounted for in these numbers, in part because community testing results are not routinely available to hospitals.

#### b) “Enhanced” nosocomial classification

Admission screening (within 24 hours) was performed in 916/1104 (83%) of admissions. All 51 definite cases had prior-negative PCRs earlier in the same admission, providing additional support for nosocomial acquisition. 56/58 (97%) probable cases had prior-negative PCRs. Although described as indeterminate, 53/79 (67%) of those diagnosed 3-7 days after admission had ≥1 prior negative sample (Table 2). Indeterminate cases with a prior-negative PCR were diagnosed later during admission (median day 5, range 3-7), than those without a prior-negative (median day 3.5, range 3-7; p=0.001). Therefore the greatest uncertainty around nosocomial acquisition exists for the 26 indeterminate cases without prior-negative PCRs and relatively short intervals between admission and first positive PCR. By definition, those without a prior-negative PCR had no prior PCR tests obtained in the same hospital admission, i.e. admission testing was not done.

**Table 2:** Epidemiological and genomic evidence for nosocomial acquisition of SARS-CoV-2 based on classification of 803 PCR-positive hospital in-patients.

|  | Definite nosocomial |  | Probable nosocomial |  | Indeterminate |  | Community-onset |  |
| --- | --- | --- | --- | --- | --- | --- | --- | --- |
| Epidemiological evidence |  |  |  |  |  |  |  |  |
| Standard epidemiological classification* | 51 |  | 58 |  | 79 |  | 615 |  |
| Enhanced epidemiological classification provides support for nosocomial acquisition (n,%)** | Yes | No | Yes | No | Yes | No | Yes | No |
|  | 51 (100%) | 0 | 56 (97%) | 2 (3%) | 53 (67%) | 26 (33%) | 89 (14%) | 526 (86%) |
| Donor/recipient closest epidemiological link identified |  |  |  |  |  |  |  |  |
| Ward contact | 46 | 0 | 53 | 2 | 51 | 24 | 69 | 0 |
| Hospital contact | 5 | 0 | 3 | 0 | 2 | 2 | 20 | 0 |
| Combined epidemiological and genomic evidence |  |  |  |  |  |  |  |  |
| Recipient and one or more epidemiological donor(s) sequenced (n,%)^ |  |  |  |  |  |  |  |  |
| Ward contact | 25/46 (54%) | 0 | 37/53 (70%) | 1/2 (50%) | 29/51 (57%) | 15/24 (63%) | 37/69 (54%) | 0 |
| Hospital contact | 1/5 (20%) | 0 | 2/3 (67%) | 0 | 1/2 (50%) | 1/2 (50%) | 5/20 (25%) | 0 |
| Genomic linkage confirmed (n,%)^^ |  |  |  |  |  |  |  |  |
| Ward contact | 17/25 (68%) | 0 | 32/37 (86%) | 1/1 (100%) | 22/29 (76%) | 6/15 (40%) | 17/37 (46%) | 0 |
| Hospital contact | 1/1 (100%) | 0 | 1/2 (50%) | 0 | 1/1 (100%) | 0/1 (0%) | 0/5 (0%) | 0 |

\* Nosocomial classification according to national Public Health England definition
\*\*Enhanced epidemiological classification for nosocomial acquisition, using prior negative PCR in the same admission, or previous hospital admission during the incubation period to provide additional support for nosocomial acquisition. Figures demonstrate number and % of nosocomial category.
^ Figures presented as number sequenced over total number of recipients with one or more ward or hospital contacts identified
^^ Figures presented as number genomically linked over total number of recipients sequenced, for ward contacts or hospital contacts

Amongst the 615 community-onset cases, a retrospective look-back at the 14 days prior to SARS-CoV-2 diagnosis, revealed 89/803 (11%) had prior hospital admissions during which SARS-CoV-2 infection could have been acquired, and 69/89 (78%) of these had ward contact with a plausible donor during that admission, suggesting reporting definitions based only on current admissions under-estimate the extent of nosocomial infection.

Similar proportions of epidemiologically plausible donors were identified for 46/51 (90%) definite cases, 55/58 (95%) probable cases and 75/79 (95%) indeterminate cases (exact p=0.52). The proportion of cases with plausible donors identified did not differ in probable or indeterminate nosocomial cases with/without prior negative PCRs (p>0.90 and p=0.59 respectively).

#### c) Epidemiological outbreak identification

Applying an epidemiological outbreak definition considering all ward overlaps led to the identification of 3 outbreaks, the largest containing over 700 individuals, highlighting that it is an unworkable definition when inpatient prevalence is high. Therefore, to more closely replicate IPC practice and provide more interpretable data, the definition of an epidemiological outbreak was restricted to only include ward overlaps with patients and staff on the ward of nosocomial diagnosis. A total of 246 individual infections (46 definite, 53 probable, 56 indeterminate, 7 community-onset and 84 staff), occurred in one of 25 outbreaks on 24 different wards. The median outbreak size was 8 (IQR 3-12, range 2-32); 99/109 (91%) definite and probable cases occurred in outbreaks.

### 3) Can genomics help to confirm/exclude nosocomial acquisition?

#### a) Genomics provides confirmatory evidence of nosocomial acquisition for most nosocomial cases

Of the epidemiologically identified nosocomial acquisitions with plausible ward donors, 107/176 (61%) samples were successfully sequenced alongside ≥1 plausible donor samples. Genomic support for nosocomial acquisition was found for 78/107 (73%) sequenced infections (17/25 (68%) definite, 33/38 (87%) probable and 28/44 (64%) indeterminate). Three further individuals with nosocomial infection, but without ward contacts, were genomically linked to a hospital-level contact (Table 2), i.e. plausibly representing transmission via an undiagnosed/unsequenced intermediate, staff contact in communal areas, staff cross-covering multiple wards or through environmental contamination.

#### b) Genomics improves precision for cases without prior-negative PCR results

Genomics helped clarify uncertainty around cases without a prior-negative PCR (Table 2). In the probable group, two individuals lacked a prior-negative PCR; one was sequenced alongside a donor and confirmed as genomically-linked (≤1 SNP), and therefore likely nosocomially-acquired. In contrast, in the indeterminate group, 26 individuals lacked a prior-negative PCR. 15/24 (63%) were sequenced alongside ≥1 potential donors; only 6/15 (40%) were genomically-linked. Hence absence of a prior-negative PCR in the indeterminate group was associated with a lower likelihood of nosocomial-acquisition, but sequencing did support some of these infections having a nosocomial source.

#### c) Genomics can confirm nosocomial acquisition in “community-onset” cases

Of the 69 individuals with community-onset infection with a prior hospital admission and a plausible donor, 37 were sequenced alongside ≥1 plausible donor(s). 17/37 were genomically-linked, indicating 17 additional infections previously categorised as “community-associated” were plausibly nosocomially acquired (Table 2).

#### d) Undiagnosed/unsequenced individuals limit utility of genomic data

Amongst the 116 nosocomial cases sequenced, 13 (11%) were genetically-linked to ≥1 other case within 0-1 SNPs but with no documented ward or hospital contact (either patient or staff). These may represent community acquisition in the case of indeterminate cases, but may also be due to undiagnosed/unsequenced individuals providing the missing epidemiological link, e.g. due to incomplete admission and ward-based patient screening and undiagnosed staff cases.

Genomic data was unable to provide confirmation of nosocomial acquisition for 22/116 (19%) of sequenced nosocomial cases. Although we can conclude that these cases were not linked to any of the other cases sequenced, we cannot use this information to exclude nosocomial acquisition from undiagnosed/unsequenced individuals, due to incomplete sampling/sequencing.

### 4) Can genomics improve understanding of transmission patterns in hospitals?

#### a) Epidemiological outbreaks often contain multiple genomically-distinct introductions

Genomic data were used to refine the epidemiologically-defined outbreaks (Figure 2B). Of the 246 staff and patients in an epidemiological outbreak, 171 were sequenced (26/46 definite, 37/53 probable, 33/56 indeterminate patients, 2/7 community-onset and 73/84 staff); 21 of the 25 epidemiological outbreaks had ≥2 members sequenced and are described further. One epidemiological outbreak was confirmed genetically as a single outbreak (each case within 0-1 SNPs of another) and in 7 ‘outbreaks’ no individuals were genetically linked (all ≥2 SNPs from all other cases). In the remaining 13 epidemiological outbreaks there was a mix of genetically-linked and unlinked cases; 9 consisted of a single genetically-distinct outbreak in addition to unlinked cases, 3 had two distinct genomic outbreaks and one ward had evidence of 3 genomic outbreaks. Overall 116/171 sequenced cases from epidemiologically defined outbreaks were confirmed to be in a genomically-supported outbreak (17/26 (65%) definite, 34/37 (92%) probable, 26/33 (79%) indeterminate, 1/2 (50%) community-onset and 38/73 (52%) staff). This highlights that epidemiological investigation may overestimate the size of outbreaks, which often occur alongside genetically-distinct introductions.

**Figure 2:**
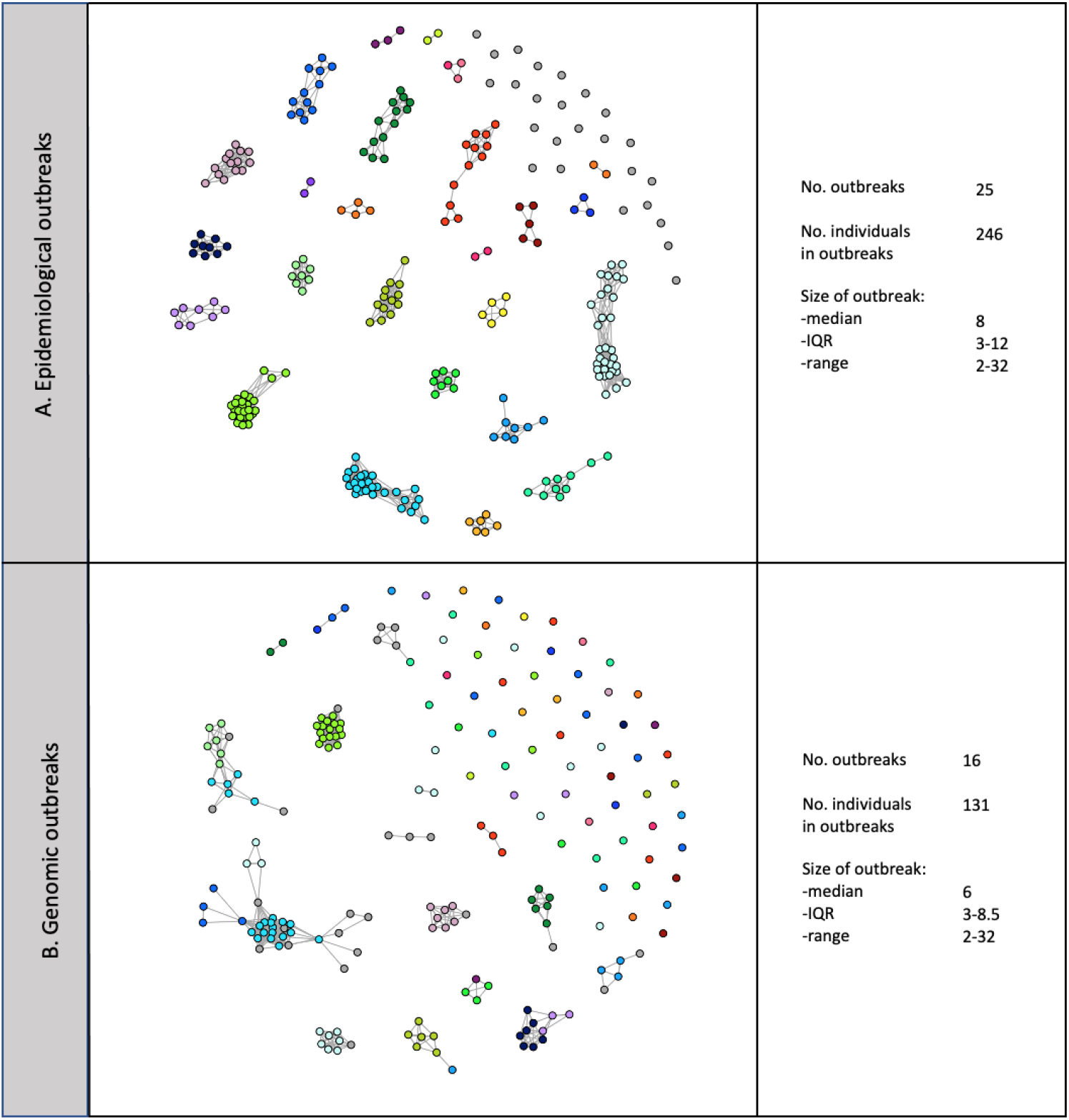
Outbreaks containing at least one definite or probable nosocomial case. A) Using epidemiological data alone (nodes are linked purely using ward-based contacts) isolated grey nodes indicate individuals in a genomic but not epidemiologically-defined outbreak, B) Using both epidemiological data and genomic data (nodes are linked both epidemiologically and genomically), isolated nodes indicate individuals in an epidemiological but not genomic outbreak. Each node represents an individual, all individuals in an epidemiological or genomic outbreak are shown in panel A with the sequenced subset in panel B. Node colours indicate the epidemiological group, grey nodes were not assigned to an epidemiological group. Lines indicate ward contact within an outbreak, line length is insignifiant. This demonstrates that epidemiological outbreaks consist of multiple genomic outbreaks and individual introductions, and conversely genomic outbreaks span multiple wards/epidemiological outbreaks. 69/176 (39%) nosocomial cases were not sequenced.

#### b) Most cases do not lead to onwards hospital transmission

Considering the cohort as a whole, rather than just those in an epidemiological outbreak as above, of the 764 individuals with samples sequenced, 200 were placed in one of 43 genomic clusters on the basis of being linked to at least one other case within 0-1 SNPs and 564 were singletons. Therefore during the period of study, SARS-CoV-2 was introduced to OUH on at least 607 occasions, with evidence of onward transmission in 43 clusters (7% of introductions) (Figure 3). The median cluster size was 2 (range 2-32). Of the 43 clusters, 17 contained both staff and patients, 16 patients only and 10 staff only. 16/43 genomic clusters were classified as genomic outbreaks (i.e. contained ≥1 definite/probable nosocomial case). Compared to the epidemiological estimate that 91% of nosocomially-acquired cases were linked to outbreaks, combining epidemiological and genomic data suggested 52/69 (75%) of all sequenced definite/probable nosocomial cases occurred in one of 16 genomic outbreaks, whereas 26% occurred as genomic singletons, ≥2 SNPs from any other case.

**Figure 3:**
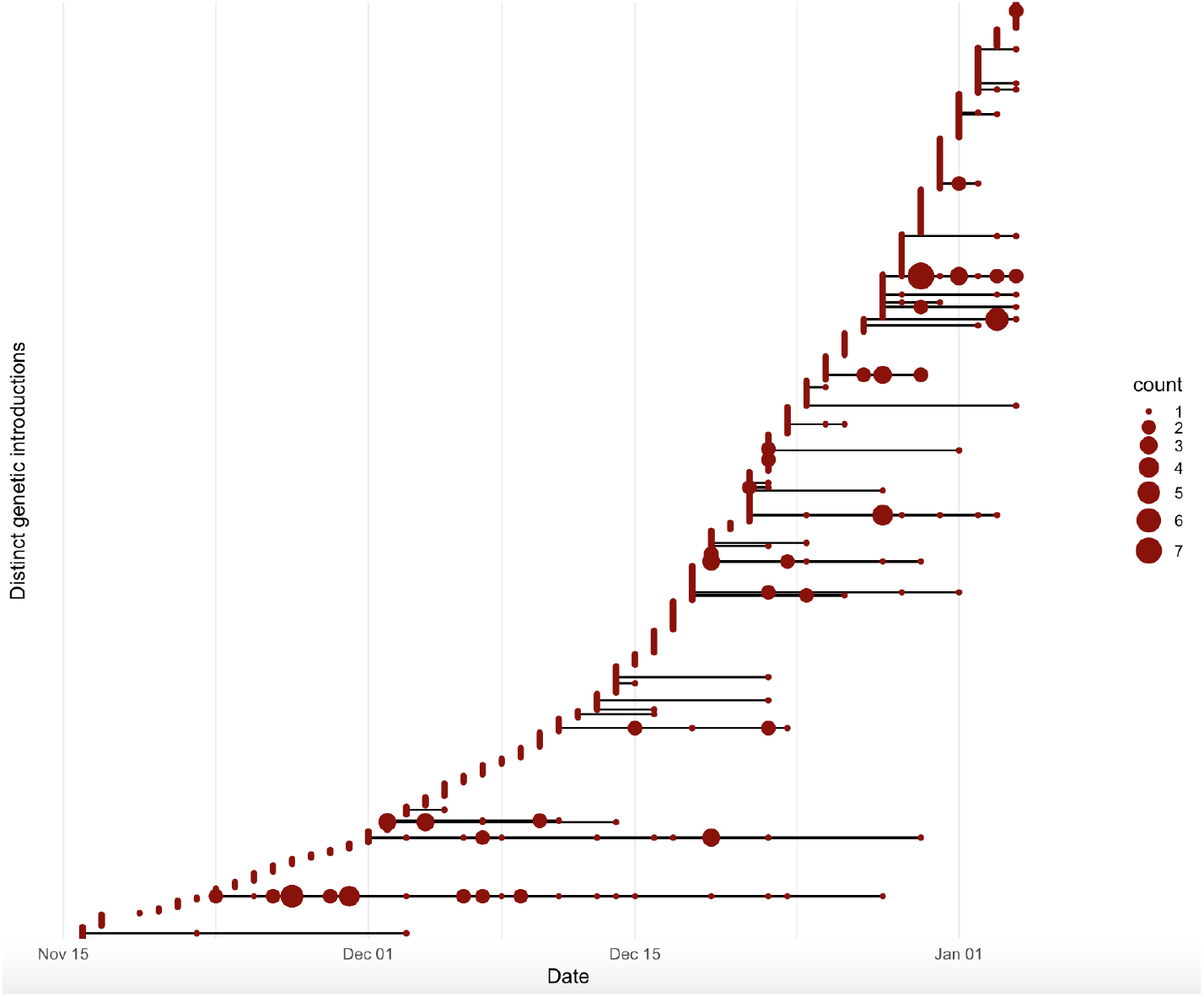
Timing and size of SARS-CoV-2 genetic clusters. SARS-CoV-2 was introduced to OUH on at least 607 occasions, with evidence of onward transmission on 43 occasions. Isolates >1 SNP from any previous sample were defined as distinct introductions and are plotted on separate horizontal lines, according to the date of the first positive sample in OUH for each individual. The size of the red dot indicates number of individuals diagnosed on each day.

Use of hospital-level ward data, accounting for all patient moves before and after testing PCR positive, led to identification of an unfeasibly large epidemiological outbreak of over 700 individuals. However, using these data in combination with WGS provides a more plausible identification of 15 additional individuals linked to outbreaks, who were missed by ward-based application of the outbreak definition due to patient ward moves during their incubation period, highlighting that outbreaks can span multiple wards.

Only 7/25 epidemiologically-defined outbreaks started with a known community-onset case, 2/7 were successfully sequenced, and only 1 confirmed to be genetically-related to subsequent cases within 0-1 SNPS. Despite the partial sequencing of community-onset cases, these data are consistent with limited direct patient-patient spread from known community-onset SARS-CoV-2 infected patients. Approximately two-thirds of staff infections were genetically distinct in this dataset, with 170/261 (65%) >1 SNP different to all other cases, across 90 work locations. Although these cases occurred on wards with existing outbreaks, they were more common in areas with transient patient contact e.g. outpatient areas and dialysis units.

The distribution of genomic clusters differed by hospital site; consistent with the extent of exposure to COVID-19 admissions; only isolated/single cases occurred at hospital “D” and only two clusters observed in hospital “C”(one staff pair and one trio containing 1 staff member and 2 patients). In contrast, hospitals “A” and “B” saw multiple larger clusters (notably, the proportion of cases sequenced was the same across all sites). In addition to differences in COVID-19 case load/infectious pressure, other factors may have played a role, such as: patient pathways including co-location of non-COVID-19 and COVID-19 cohort wards, estates/facilities, including number of patient side rooms and ventilation, differences in staff mobility between COVID-19 and non-COVID-19 wards, staff facilities (communal/break areas) and adherence to social distancing.

#### c) Two patterns of nosocomial acquisition seen

Broadly two patterns of nosocomial acquisition were seen; patterns are shown on a representative example phylogeny in Figure 4.

**Figure 4:**
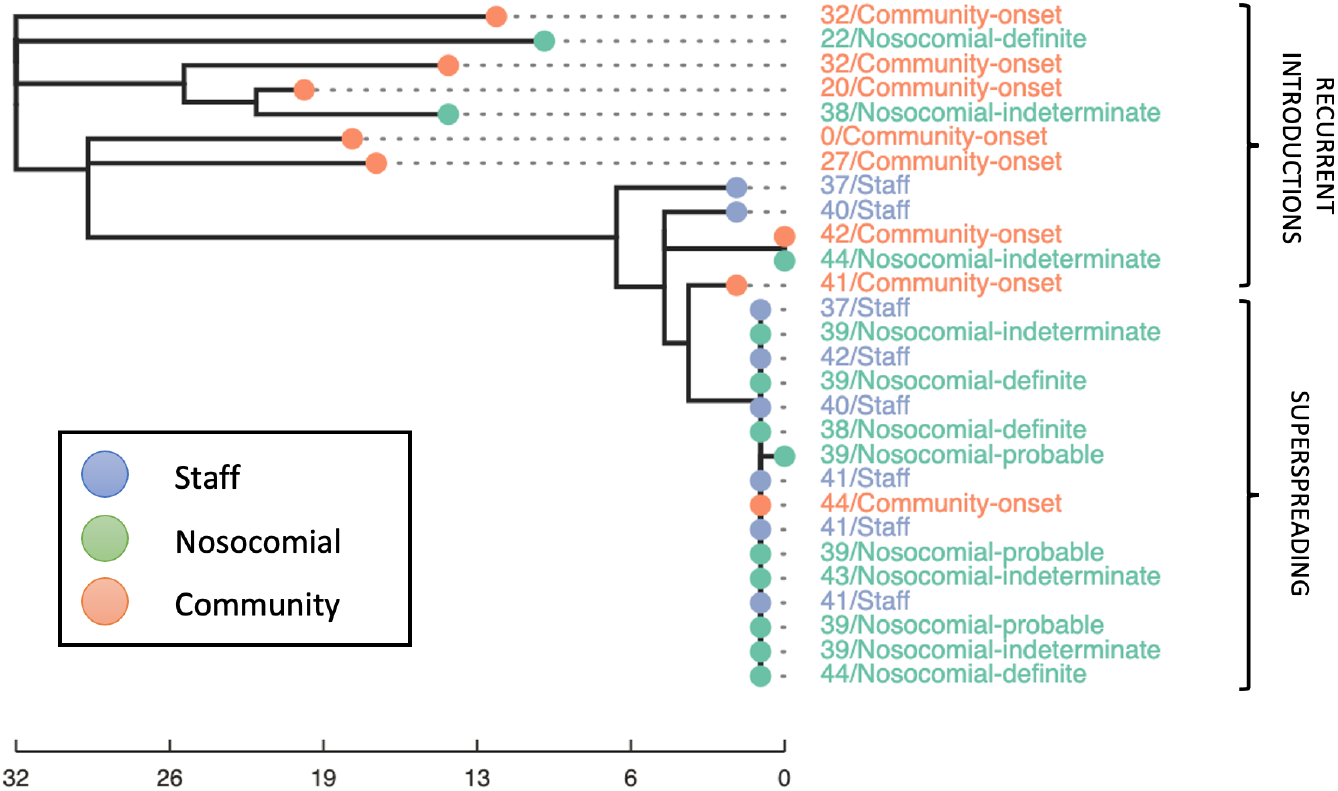
Example phylogeny demonstrating a superspreading event and recurrent introductions on a single ward at hospital “B”. The node and label colour indicates broad epidemiological classification (community-onset, nosocomial, staff). The tip label gives the day of the outbreak the individual tested positive followed by the full epidemiological classification. The scale bar represents SNP distance. All cases were classified as part of the same epidemiological outbreak, however WGS reveals multiple introductions. The “community-onset” case diagnosed on day 44 of the outbreak, had a previous hospital admission with exposure on this ward during a superspreading event.

##### i) Superspreading events

These are characterised by a rapid accumulation of multiple cases within 1-2 serial intervals, e.g. >5 cases within a 7-10 day period, implying multiple transmissions per serial interval. 8 such events occurred during this study, 7 at hospital “A”, and one at hospital “B”, typically occurring on non-COVID-19 wards, with open bays, involving both staff and patients, and in specialties with patients highly dependent on nursing care (e.g. trauma, acute medicine, neurology). The median outbreak size was 9 (range 7-32) and median duration 12 days (range 7-35 days). Although infrequent, these 8 superspreading events accounted for 80% of cases linked to a genomic outbreak.

With a serial interval of 5 days, some outbreaks may represent exposure to a single superspreading infection, but most are subsequently propagated amongst staff/patients. Incomplete sampling and asymptomatic individuals without symptom onset dates prevents confident identification of the source of each outbreak, however, in two clusters, staff cases preceded patient cases, so staff could have acted as an index events. In the remaining six outbreaks, there were no cases diagnosed prior to the first definite/probable nosocomial case to act as a plausible index, therefore the outbreak was likely seeded by an undiagnosed or unsequenced patient/staff/visitor. No outbreaks were seeded by direct patient-patient transmission from known positive patients, however we cannot exclude a non-sequenced cross-covering staff member providing the missing epidemiological link, by acquiring infection from a known positive patient and seeding an outbreak on a non-COVID ward.

##### ii) Recurrent introductions

These are characterised by slow “rumbling” accumulation of nosocomial and staff cases on a ward, on both non-COVID and mixed wards with side rooms accommodating COVID and non-COVID patients (Figure 4). They consist of multiple introductions of distinct viral variants over a more prolonged period of time, giving the appearance of a slowly progressing outbreak, but with no, or minimal, onward transmission within the unit. Genomic data is required to distinguish recurrent introductions from genomic outbreaks.

Recurrent introductions involving one or more definite/probable case occurred on 6 different wards across hospitals “A”, “B” and “D”. Each mimicked an outbreak with between 3-6 staff and nosocomial cases occurring on the ward over 2-6 week periods, however all were genetically distinct introductions.

## Discussion

In this retrospective cohort study of healthcare-associated SARS-CoV-2 transmission using combined epidemiological and sequencing data we make several key findings that challenge current surveillance definitions and reveal most nosocomial transmission occurs from a relatively limited number of highly infectious individuals.

Our findings suggest that the majority of cases occurring after >7 days in hospital are nosocomially acquired, for example 107/109 probable/definite cases had ≥1 prior-negative PCR test in the same admission, and most had plausible ward-based sources for their infection. However, surveillance definitions identifying probable/definite nosocomially-acquired cases (on the basis of prior hospital stays of >7 days) likely under-estimate the extent of acquisition in hospital. In the UK, nationally reported nosocomial figures exclude indeterminate cases (diagnosed on day 3-7 of their hospital stay); however several of these cases in our study had prior-negative PCR tests during the same admission, plausible exposure to infectious patients, and genomic-linkage with other cases, all supporting acquisition in hospital, particularly for cases diagnosed on days 5-7. Furthermore, surveillance definitions considering only the current hospital stay, as in the UK, do not capture nosocomial acquisition during a recent prior hospital stay, for which we found both epidemiological and genomic evidence. Consideration should be given to revising surveillance definitions to account for prior-negative tests and infections diagnosed <7 days into admission.

Consistent with defining most cases within 2 days of admission as community-acquired, genomics demonstrated most cases in staff and patients are genomically-distinct from all others in the hospital; there were 607 genomic clusters within the 764 samples sequenced. This is similar to WGS-based findings in other healthcare-associated infections over the last decade[28,29]. However, in contrast to other nosocomial infections, we found evidence that most nosocomial acquisition occurs in explosive superspreading events, with clusters of genomically-related cases occurring in short time periods, as observed by others for SARS-CoV-2 in both community and hospital settings [9,30–33].

WGS added most value when investigating outbreaks during periods of high SARS-CoV-2 prevalence, given high rates of ward-based contact with infected patients. The majority of epidemiologically-defined outbreaks consisted of multiple genomic introductions with some smaller genomic clusters. The role of staff in outbreaks is overestimated from epidemiological data alone, with genomics confirming only 52% of staff epidemiologically placed in an outbreak were genomically-linked, and the majority of sequenced staff cases were genomic singletons (≥2 SNPs from any other case). Additionally, in hospitals not routinely admitting COVID-19 patients, rates of transmission were low, suggesting that isolated acquisition from staff is relatively uncommon, and that transmission requires a ‘perfect storm’ of mixed COVID and non-COVID wards, emergency admissions and dependent patients accommodated in bays.

The main limitations of genomic data were two-fold. Firstly, although epidemiological data is available for all patients, genomic data is limited by sample availability and difficulty of generating sequences at low viral loads. Here 67% of the cohort were successfully sequenced, in line with other similar hospital cohorts (20-70%)[7,11,34]. As such, genomic data does not enable nosocomial acquisition to be ruled out. Incomplete hospital sequencing datasets suffer from an ‘absence of evidence’ when attempting to exclude nosocomial acquisition, which should not be mistaken for ‘evidence of absence’ of nosocomial acquisition. This may be mitigated in the future by integrated community epidemiological and genomic datasets, and could be addressed through probabilistic inference methods that can account for missing data, or by further optimising sequencing yields. Future approaches to evaluating transmission could also consider proxy markers of infectiousness such as Ct values (reflecting viral loads).

Secondly, the rapid transmission of SARS-CoV-2 in relation to viral evolution and the short time spans of outbreaks are insufficient for substantial genetic variation to accumulate, and therefore genomic data alone is insufficient to confer linkage or resolve the ordering of transmission; a combination of epidemiological and genomic data is required. A 1 SNP cut-off for defining linkage captures the majority of cases genuinely linked to the cluster, with the compromise of including a few community-onset cases likely linked by chance. The sensitivity and specificity of this SNP threshold for defining linkage varies according to the point during the pandemic at which it is being applied, both in terms of time since the start of the pandemic (greater overall viral diversity afforded by later time points in the pandemic), and the current rate of transmission (locally reduced diversity during exponential periods of spread, such as was observed with the emergence of the alpha variant in the winter of 2020). Improvements in sensitivity and specificity to detect transmission might also be gained from considering patterns of intra-host variation[35,36]. Generally, however, joint epidemiologic and genomic analysis enable the limitations of one method to be compensated for by the strengths of the other, acknowledging both approaches are limited by undiagnosed cases.

Our data have several practical IPC implications. The small proportion of cases leading to detected onward transmission highlight that existing enhanced IPC practices are generally effective at preventing most patient-patient spread from known positive patient cases, e.g. via triaging of patients into pathways on admission and widespread diagnostic testing reducing contact between infectious/susceptible individuals. However, rates of nosocomial infection remain too high, with the highest rates in hospitals caring for both COVID-19 and non-COVID-19 patients.

When investigating nosocomial SARS-CoV-2 transmission, it is vital to consider the contributions of both patients and staff in initiating and amplifying transmission. The identification of superspreading events highlights the importance of screening of both asymptomatic patients and staff to help identify and control outbreaks early (whilst acknowledging that virus in up to half of positive staff may not genetically be part of the outbreak). Several strategies can be used and scaled according to the situation, from universal admission testing, weekly ward screening, asymptomatic staff lateral flow device testing as standard, scaling up to on-demand full-ward lateral flow and PCR screening, and more frequent regular screening if nosocomial cases are identified. If multiple cases are observed within a single serial interval, highly suggestive of a superspreading event, rapid action should be taken, which may involve temporary ward closure to mitigate secondary transmission, recognising that those recently infected have the highest viral loads[37] and are most infectious to patients and staff.[38] If resources allow, use of dedicated staff in high risk areas, and self-isolation at home for staff exposed to a high risk event, may also be appropriate. Challenges include recognising outbreaks spanning multiple areas and implementing effective testing and control measures, e.g. for patients who move between wards and staff who cross-cover multiple wards, including during nights and those contracted by outside agencies. Communication that patients discharged from a superspreading ward are at high risk for acquisition should lower the threshold for post-discharge SARS-CoV-2 screening/testing. Variations in rates of nosocomial transmission suggest screening should be prioritised on wards and in specialities with the highest risks (e.g. acute medicine, trauma, neurology in our setting). As vaccination-mediated reductions in inpatient COVID cases occur, it will be important to raise awareness that patients on low risk wards/pathways are still at risk of nosocomial acquisition, in addition to highlighting that in general outbreaks are caused by patients or staff not known to be positive.

This study demonstrates that retrospective analyses of genomic data is useful in some circumstances to guide future IPC practice, with results consistent with similar studies in the UK [7–10]. It remains to be seen whether the additional costs of generating and analysing this genomic data near real-time (<48hrs from sample to dissemination of results) are justified by additional IPC gains, or whether the rapid and rigorous application of gold standard epidemiological methods in response to fast accumulation of nosocomial PCR-based diagnoses is the key intervention. This question will be addressed by studies such as the COG-HOCI trial[39]. Regardless of WGS, there is a clear need for automated systems to rapidly assimilate epidemiological data tracking patients over space and time to allow transmissions based on locations other than ward of diagnosis to be quickly identified and fed to IPC teams.

In conclusion, epidemiological investigation can be enhanced by genomic data, to provide insights into nosocomial acquisition and outbreaks in the hospital setting, and provide practical insights to optimise IPC interventions.

## Acknowledgements

We would like to thank all OUH staff who participated in the staff testing program, and the staff and medical students who ran the program. This work uses data provided by healthcare workers and collected by the UK’s National Health Service as part of their care and support. We thank all the people of Oxfordshire who contribute to the Infections in Oxfordshire Research Database. Research Database Team: L Butcher, H Boseley, C Crichton, DW Crook, DW Eyre, O Freeman, J Gearing (community), R Harrington, K Jeffery, M Landray, A Pal, TEA Peto, TP Quan, J Robinson (community), J Sellors, B Shine, AS Walker, D Waller. Patient and Public Panel: G Blower, C Mancey, P McLoughlin, B Nichols

## Supplement

### Group authorship

#### Oxford University Hospitals microbiology laboratory (Oxford University Hospitals NHS Foundation Trust, Oxford, UK)

Anne Baby, Jasmine Bastable, Kathryn Cann, Reena Chohan, Josie Clarke, Gabriel Cogorno, Sam Cordy, Georgina Coward, David Crawford-Jones, Sean Crawley, Jack Dobson, Bronte Drummond, Laura Dunn, Caleb Edwin, Simon Evans, Mohamad Fadzillah, Jess Gentry, Sarah Hill, Laura Hobden, Nurul Huda, Gemma Innes, Scott Jarvis, Gerald Jesuthasan, Emma Jones, Anita Justice, Lizzie Kalimeris, Richard Kirton, Nakiah Lashley, Sophie Mason, Alex Mobbs, Ahila Murugathasan, Eleanor Mustoe, Gospel Ngoke, Sarah Oakley, Oliver O’Sullivan, Kimberley Odwin, Jack Oliver, Freyja Pattrick, Claudia Pereira, Simon Perry, Tom Potter, Alexander Prentice, Sophie Ramage, Athena Sanders, Kellyanne Savage, Katherine Shimell, Robin Terry, Emma Thornton, Sue Wareing, Annie Welbourne, Maddison Wheatley

#### Oxford University Hospitals Infection, Prevention and Control team (Oxford University Hospitals NHS Foundation Trust, Oxford, UK)

Gabriella D’Amato, Ruth Moroney, Gemma Pill, Lydia Rylance-Knight, Claire Sutton, Claudia Salvagno, Merline Tabirao, Sarah Wright

### Supplementary methods

#### Summary of PHE IPC and testing recommendations

– Patient pathways - establish separation of patient pathways and staff flow to minimise contact between pathways. Three pathways identified:
  – high risk - confirmed COVID-19 positive by a SARS-CoV-2 PCR test or are symptomatic and suspected to have COVID-19 (awaiting result)
  – medium risk - waiting for SARS-CoV2 PCR test result and who have no symptoms of COVID-19 and individuals who are asymptomatic with COVID-19 contact/exposure identified
  – low risk – triaged/tested (negative)/clinically assessed with no symptoms or known recent COVID-19 contact/exposure
– Ensure that hygiene facilities, IPC measures and messaging are available for all patients/individuals, staff and visitors to minimize COVID-19 transmission. For example universal masking, physical distancing, hand hygiene, increasing frequency of equipment decontamination and improving ventilation (aerosol generating procedures should only be carried out when essential with airborne PPE precautions).
– Visiting restrictions dependent on patient pathway
– Inpatient testing:
  – all patients at emergency admission, whether or not they have symptoms;
  – those with symptoms of COVID-19 after admission;
  – for those who test negative upon admission, a further single re-test should be conducted between 5-7 days after admission;
  – test all patients on discharge to other care settings, including to care homes or hospices;
  – elective patient testing prior to admission.
– Staff testing
  – all staff with symptoms (or the index case if a household member)
  – non-symptomatic staff (in addition to all patients and symptomatic staff) working in situations where there is an untoward incident or outbreak or high prevalence.

#### Study design, setting and participants

Patients received an admission PCR test regardless of symptoms, with weekly asymptomatic ward screening thereafter, increasing to twice weekly from 01-December-2020, with symptomatic testing as required. Symptomatic and twice monthly asymptomatic staff testing programmes were available throughout the study period as previously described,[5] and voluntary twice weekly lateral flow device testing was introduced for staff from 23-November-2020.[40]

Of the 2773 PCRs performed on this cohort during the study period, 2553 (92%) were performed using the Thermo Fisher TaqPath assay (targeting S and N genes, and ORF1ab; Thermo Fisher, Abingdon, UK), 78 on Altona RealStar (targeting E and S genes; Altona Diagnostics, Liverpool, UK), 64 on Abbott RealTime (targeting RdRp and N genes; Abbott, Maidenhead, UK), 59 on Cepheid Xpert® Xpress SARS-CoV-2 (targeting N2 and E; Cepheid, California, USA) and 19 on BioFire® Respiratory 2.1 (RP2.1) panel with SARS-CoV-2 (targeting ORF1ab and ORF8; Biofire diagnostics, Utah, USA).

A result was considered positive on the Thermo Fisher platform if 2 out of 3 gene targets in addition to the positive control were detected (with Ct ≤32 for one target and ≤33 for another). Samples with all targets with Ct >37 or not detected were considered negative, other results were considered indeterminate.

#### Sequencing methods and quality control metrics

Samples were sequenced using a multiplex PCR-based approach with the ARTIC LoCost protocol and v3 primers[14] using R9.4.1 flow cells (Oxford Nanopore Technologies, Oxford, UK). Consensus sequences were generated using the ARTIC fieldbioinformatics v1.2.1 Nanopolish workflow. [41] All samples underwent quality control, requiring >90% consensus genome coverage at ≥20x depth. Repeat isolates from the same individual were not routinely sequenced; if multiple samples per individual were stored, the lowest Ct sample was selected for sequencing. For 86 cases where duplicate samples were sequenced for an individual, the sample with highest percentage genome coverage was selected for analysis.

A total of 764 samples passed quality control. The median % genome coverage was 98% (IQR 96 - 99%), the median depth of coverage was x915 (IQR 633 - 1346).

#### Sequence comparison

A multiple sequence alignment (MSA) of 764 sequences based on Wuhan-Hu-1 was constructed using MAFFT parameters ‘--auto --6merpair --addfragments’. Phylogenetic reconstruction from the MSA was performed with IQTree parameters ‘-m GTR+G -blmin 1e-9’. The phylogeny was rooted with Wuhan-Hu-1 and branch lengths were SNP-scaled.

#### Defining genetic relationships

The estimated evolutionary rate for SARS-CoV-2 is 1.1×10^−3^ substitutions/site/year, which equates to 2-3 substitutions per genome per month.[42] This evolutionary rate is slow compared to the rapid transmission of cases in the community and in hospital, with a serial interval of approximately 5 days.[18] This leads to the possibility that two people may be infected with identical viruses by chance, rather than due to direct person-to-person transmission. Using a cumulative Poisson distribution, with approximations of one mutation every 2 weeks and a serial interval of 5 days, there is 70% chance of no new SNPs per transmission, 95% of 0-1 SNPs, and 99% of 0-2 SNPs, therefore the higher the SNP threshold set for considering transmission, the more true transmissions are captured. However, the extent of population-level diversity also needs to be considered, i.e. the probability of two randomly chosen sequences being within 0, 0-1, or 0-2 SNPs within this data set was 0.9%, 2.1% and 3.5% respectively. Therefore a plausible genetic relationship was defined as ≤1 SNP between two samples in this study, an association close enough to support transmission whilst minimising over-calling of linkage in non-nosocomial cases.

## Supplementary tables and figures

**Supplementary table 1:** Sequencing pass rates by nosocomial classification and Thermofisher Ct value.

|  | Total | Pass |  |  | Fail |  |  | Not available |  |  |
| --- | --- | --- | --- | --- | --- | --- | --- | --- | --- | --- |
|  |  | TF Ct <28 | TF Ct >28 | Non TF | TF Ct <28 | TF Ct >28 | Non TF | TF Ct <28 | TF Ct >28 | Non TF |
| **Definite** | 51 | 24 | 3 | 0 | 4 | 16 | 0 | 2 | 0 | 2 |
| **Probable** | 58 | 33 | 9 | 0 | 2 | 11 | 0 | 1 | 0 | 2 |
| **Indeterminate** | 79 | 39 | 8 | 0 | 6 | 22 | 1 | 1 | 2 | 0 |
| **Community-onset** | 615 | 361 | 26 | 0 | 56 | 93 | 0 | 21 | 35 | 23 |
| **Staff** | 329 | 238 | 20 | 3 | 9 | 44 | 2 | 1 | 1 | 11 |

TF Ct <28 = Ct value of sequenced sample was less than 28
TF Ct > 28 = Ct value of sequenced sample was more than 28
Non TF = indicates the PCR was not performed on the Thermo Fisher platform (to allow comparison, only Thermo Fisher Ct values are reported).

**Supplementary table 2:** Demographics of sequenced vs non-sequenced inpatients.

|  | All patients | Sequenced | Not sequenced |
| --- | --- | --- | --- |
| **Total number** | 803 | 503 (63%) | 300 (37%) |
| **Sex** |  |  |  |
| Male (n; %) | 454 (57%) | 292 (58%) | 162 (54%) |
| Female (n; %) | 349 (43%) | 211 (42%) | 138 (46%) |
| **Age (median; IQR)** | 67 (49-81) | 69 (51-81) | 63 (43-80) |
| **Lowest Ct (median; IQR)** | 20 (14-27) | 17 (14-21) | 28 (22-32) |
| **Length of stay before first SARS-CoV-2 positive PCR, days (median; IQR)** | 1 (1-3) | 1(1-2) | 1 (1-5) |
| **Length of stay after first SARS-CoV-2 positive PCR, days (median; IQR)** | 6 (2-13) | 7 (2-14) | 6 (2-11) |
| **28 day mortality (n; %)** | 129 (16%) | 97 (19%) | 32 (11%) |

**Supplementary figure 1:**
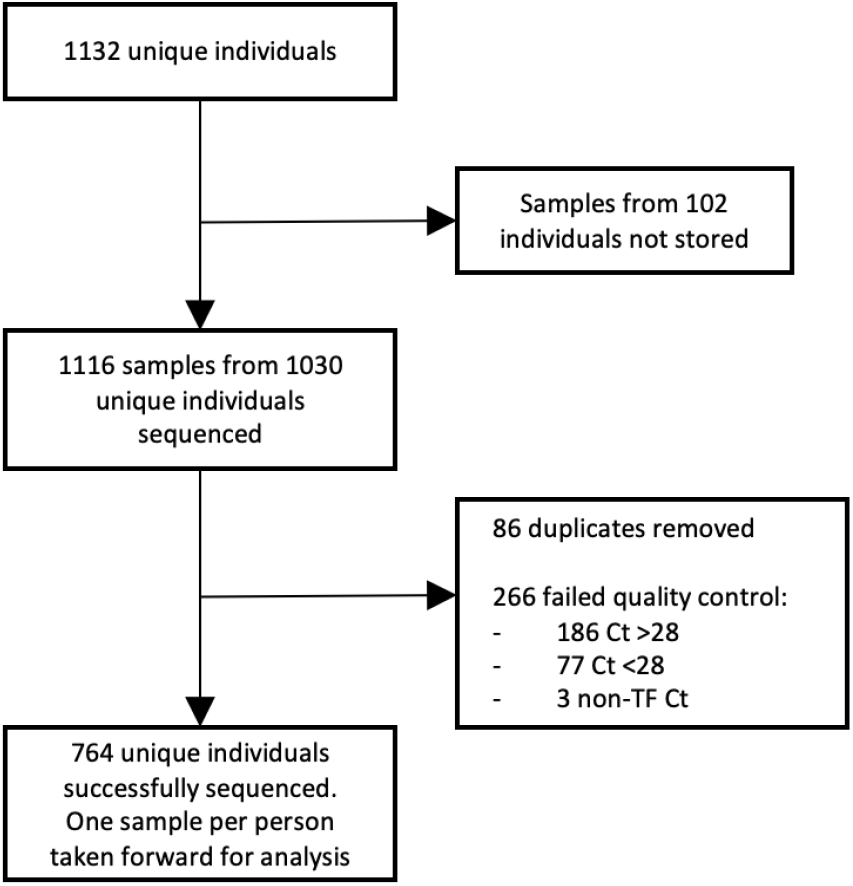
Flow diagram showing numbers of samples available for sequencing and being successfully sequenced. Where duplicate samples from an individual were sequenced, the sample with highest percentage SARS-CoV-2 genome coverage was selected for analysis. TF = Thermofisher (only Ct values for the most commonly used Thermofisher PCR platform are reported).

